# Ramadan and Kidney disease (RaK) risk assessment tool. Potential Risk Calculator for Evaluating the Risk of Ramadan Fasting In Chronic Kidney Disease patients

**DOI:** 10.1101/2025.02.20.25322592

**Authors:** Yousef Boobes, Nico Nagelkerke, Fatima Alkindi, Bachar Afandi, Hanan Abdelbaki, Esraa Abdelkhalig Mahmoud, Fatima Yousef Almeqbaali, Mahra Ibrahim Alyahyaei, Jamal Eldin Sahyouni, Aysha Abdulla Al-Kuwaiti, Abdallah Abukhater, Aamna Ghanim Albloushi, Amal Mohammed Alshehyari, Sara Fadel Al Shukri, Rudina Mubarak Alketbi, Maitha Sultan Alaryani, Fatima Ali Alketbi, Amna Mohammed Al Ahbabi, Toqa Abdel Rahman Fahmawee, Waad Mashaal Alantall, Noor Abdullah Yahya, Latifa Mohammad Baynouna AlKetbi

## Abstract

**Background:** Fasting during Ramadan has been found safe for most chronic kidney diseases (CKD) patients but may pose risks for some. CKD patients who choose to fast require careful attention from healthcare providers. However, there is a lack of studies evaluating the impact of Ramadan fasting on various kidney diseases. The Ramadan and Kidney Disease (RaK) working group has developed a risk stratification tool to categorize CKD patients intending to fast into low, moderate, and high-risk groups. We propose a Ramadan fasting calculator incorporating key risk factors to help guide fasting decisions in CKD patients.

**Methods:** A prospective cohort study was conducted at Ambulatory Healthcare Services from March 2024 to August 2024. The Electronic Medical Records (EMRs) of 220 patients were reviewed, supplemented by teleinterviews conducted before and after Ramadan. The primary outcomes assessed were the Ramadan fasting experience and the occurrence of significant adverse events (SAEs).

**Results:** Among the 220 participants, 71.4% completed all 30 days of fasting during Ramadan, 7.3% began fasting but had to break it, and 21.4% did not fast. A total of 39 significant adverse events (SAEs) were recorded. This sample’s mean and median RaK scores were 9.78 and 9.0, respectively. Distribution of RaK scores was as follows: 55.5% had a score of 9 or lower, 24.5% had scores between 10 and 13, and 18.6% had scores above 13.

Ordinal regression (proportional odds model) was used to predict fasting status (categorized as 0 = no fasting, 1 = partial fasting, 2 = completed fasting). The RaK score was the only significant predictor, with an estimate of −0.159 (95% CI: −0.264 to −0.053), p = 0.003 (Figure 2). The area under the curve (AUC) for predicting the likelihood of attempting fasting was 0.715 (95% CI: 0.635–0.794), with an optimal cutoff point of 9.5.

Regarding SAE prediction using logistic regression, a higher RaK score was significantly associated with increased SAE incidence (B = 0.253, OR: 1.036–1.599, p = 0.023). Additionally, non-UAE nationality and frailty were also significantly associated with a higher risk of SAEs.

**Conclusion:** The RaK risk assessment tool was found to be a valid predictor of fasting status and significant adverse events (SAEs) in CKD patients. This is the first validated risk assessment tool designed for CKD patients intending to fast during Ramadan. Further studies are needed to refine its applicability and provide more tailored guidance for specific subgroups of kidney patients.

**Key learning points:** What was known:

Research on Pre-Ramadan risk assessment in relation to Ramadan fasting by kidney disease patients is lacking.

This study adds:

This is the first study to validate the RaK assessment tool developed based on multidisciplinary experts’ consensus.

The tool was effective in identifying patients at high risk of developing adverse events during fasting and in identifying patients who can fast safely.

Potential impact:

The RaK assessment tool is recommended to be included in the pre-Ramadan assessment of patients with kidney diseases.

## Introduction

Fasting during the holy month of Ramadan is one of the five pillars of Islam. Muslims must abstain from eating or drinking from dawn until sunset. Ramadan Fasting (RF) is reported to be safe for most kidney disease patients and may even contribute to moderate improvements in kidney function (1). However, several factors can increase the risk of adverse health outcomes during fasting, particularly in patients with chronic kidney disease (CKD), who often have multiple comorbidities that complicate their management (2).

CKD is a significant global public health issue, affecting approximately 13% of the population worldwide. In the United Arab Emirates, the estimated prevalence of CKD in Abu Dhabi is around 8%. Given its high prevalence and associated complications, CKD represents a substantial health burden both globally and regionally (3) (4)

Muslims with sickness are exempted from fasting, but decision-making by physicians and patients is complex, as the risk of harm is probabilistic and cannot be predicted with certainty. To provide individualized recommendations for patients with CKD, an accurate assessment of the associated risks is essential. While a risk stratification tool for identifying high-risk diabetes patients has been developed and validated by the IDF-DAR, no such tool previously existed for CKD patients (5) The Ramadan and Kidney Disease Initiative (RaK) was established in May 2022 to address this gap. This initiative brought together an international panel of experts, including nephrologists, endocrinologists, and family medicine physicians with expertise in CKD and Ramadan fasting. The Ramadan and Kidney Disease Working Group developed a risk assessment and stratification tool and clinical recommendations to guide fasting decisions for CKD patients and their healthcare providers.(1) (6)

To develop these guidelines, the RaK group conducted a systematic review of studies on Ramadan fasting and its associated risks. This review led to the development of a scoring system that stratifies patients into different risk categories, aiding in counseling patients on fasting during Ramadan. Due to the limited availability of quantitative studies in this area, most of the scoring system’s development was based on expert consensus. This study is the first to quantitatively validate the RaK scoring tool among CKD patients considering Ramadan fasting, assessing its predictive value for both fasting completion and adverse event occurrence. A secondary aim is to stratify high-risk CKD patients in Abu Dhabi using the RaK tool.

## Method

This study is a prospective cohort study conducted from March to August 2024. It included patients aged 18 years or older attending Ambulatory Healthcare Services (AHS) centers who were either UAE nationals or foreign residents of the Abu Dhabi Emirate. The AHS CKD Population Health Program was implemented in 2018 at the AHS centers. The lab results for all patients at AHS with abnormal kidney function tests are reported to the AHS centers for follow-up and management. Patients who appeared in the report between October and March 2024 were included in this study, resulting in 450 eligible patients. Due to time and resources, only 330 were contacted, and 271 were reachable and agreed to participate. Only 220 accurately reported their fasting status, and those who fasted or attempted fasting were included in the analysis; 173 did not fast a single day while 47 did not participate in fasting.

Participants were called and tele-interviewed twice before and after Ramadan. Consent to participate was sought from each participant, and an EMR audit was conducted for all those agreeing to participate after each interview. The first tele-interview was in March 2024, before Ramadan. CKD patients were contacted and assessed using the RaK proposed risk assessment tool (supplementary 1). The Rak tool included age, GFR level, level of Proteinuria, presence of diabetes, glycemic control, hypertension, presence of CVD, history of a kidney transplant, history of kidney stones, medications used (CNI, Diuretics, SGLT2 I, ARB/ACE-I), country duration of fasting in hours, patient expose to temperature, physical Labor, past fasting experience and history of acute kidney injury during last 3 months. Important demographics and medical histories such as sex, nationality, health conditions, and patients’ assessments such as SBP, DBP, and BMI were also collected. Most lab results were available for the patient’s routine care, including HBA1C and renal function. Frailty assessment was done for patients over 60 using the FRAIL tool. The FRAIL tool is a validated questionnaire with five questions predicting clinical outcomes (7). The five questions were related to fatigue, resistance or climbing stairs, Ambulation (i.e., walking a couple of blocks, number of chronic illnesses, and losing weight by more than 5%. The data collection was done by family medicine residents.

### Outcome assessment

After Ramadan, additional data were collected to assess significant outcomes, such as illnesses, admissions, or any health issues that developed during fasting. Only events occurring during Ramadan were considered in this study. The surveillance started on the first day of Ramadan and continued until the end of the holy month. Any deterioration in renal function was assessed within 3 months after Ramadan.

### Data Analysis

To assess the statistical power of our study, we considered a scenario in which 25% of the patients would have a specific event (“cases”) and 50% of the “controls” would be “exposed” to a binary risk factor. Then, our sample of 220 would be able to demonstrate an odds ratio of 2.5 for the association between that risk factor and the event with 80% power and 5% significance level. Greater power can be expected for continuous risk factors. Statistical analysis was done with SPSS v29. Patients with CKD who fasted all Ramadan days or attempted to fast were included. Type I DM patients were excluded (3 patients). Fasting status and significant events were treated as dependent variables. Other variables were treated as independent variables, including the RaK risk category. Frequencies, cross-tabulation, proportional odds, logistic, and linear regression were used. C-statistics were used to test the prediction model’s performance derived from the regression analysis.

### Patient and Public Involvement

Patients or the public WERE NOT involved in the design, or conduct, or reporting, or dissemination plans of our research.

## Results

Of the 220, 71.4% completed all 30 days of fasting during Ramadan, 7.3% began/attempted to fast but had to break their fast, and 21.4% did not fast a single day. There were 39 significant adverse events, with 12 (30.8% better to use percentage of group NOT % of events) among those who did not fast, 8 (20.5%) among those attempting, and 19 (48.7%) among those who completed their fast. Admission was the main event, 30 out of the 39, a very strong indicator of severity.

Our sample included many high-risk patients: 67.3%were 60 or older were, nine patients had a history of transplant, four were on dialysis, 20.1% on insulin, 56.8% were diabetics, 7.3% experienced AKI in the last 3 months before Ramadan, 25% were frail, and 13.6% had a hospital admission within 3 months before Ramadan. Table 1 presents more characteristics of the sample.

**Table 1.**
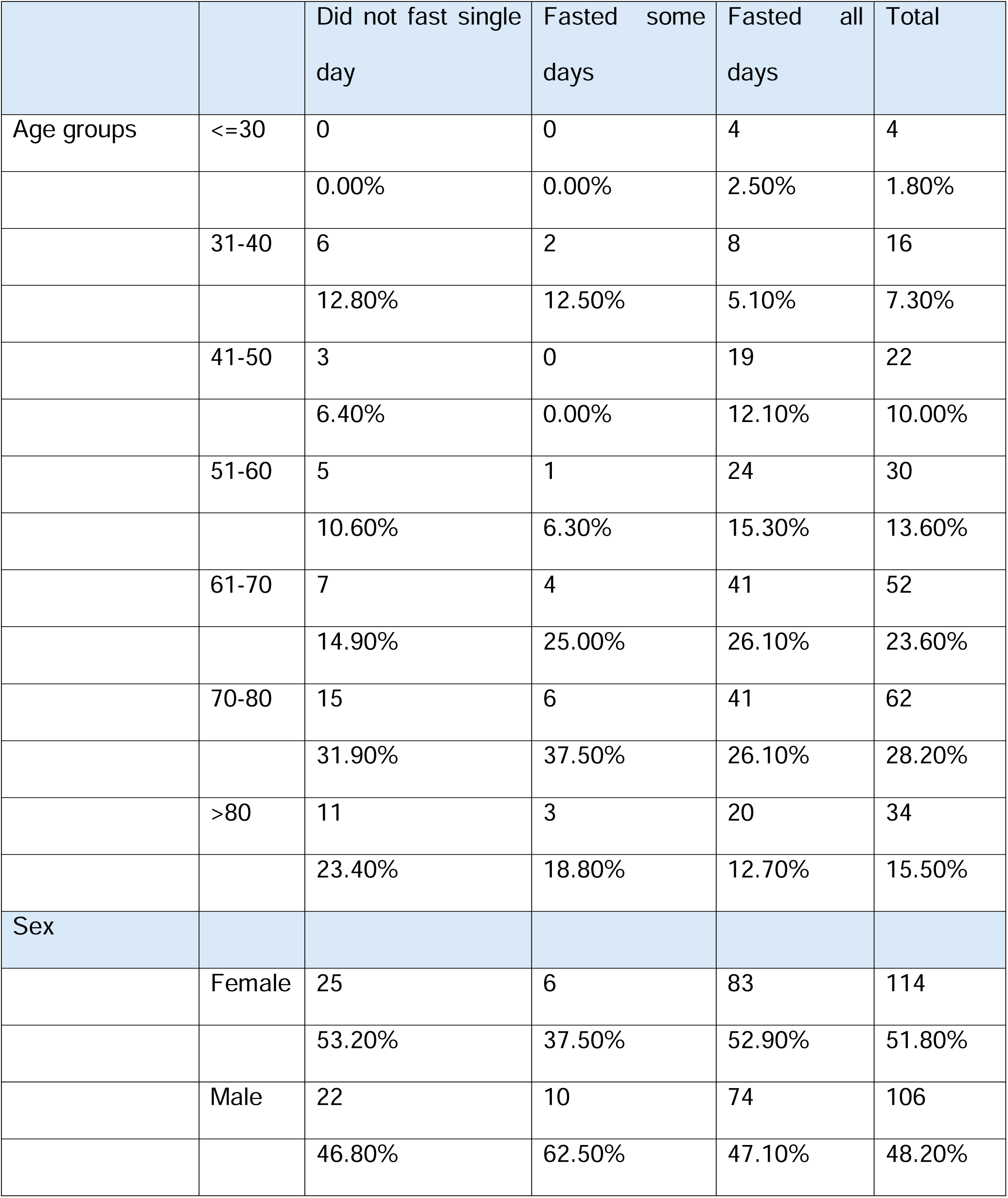
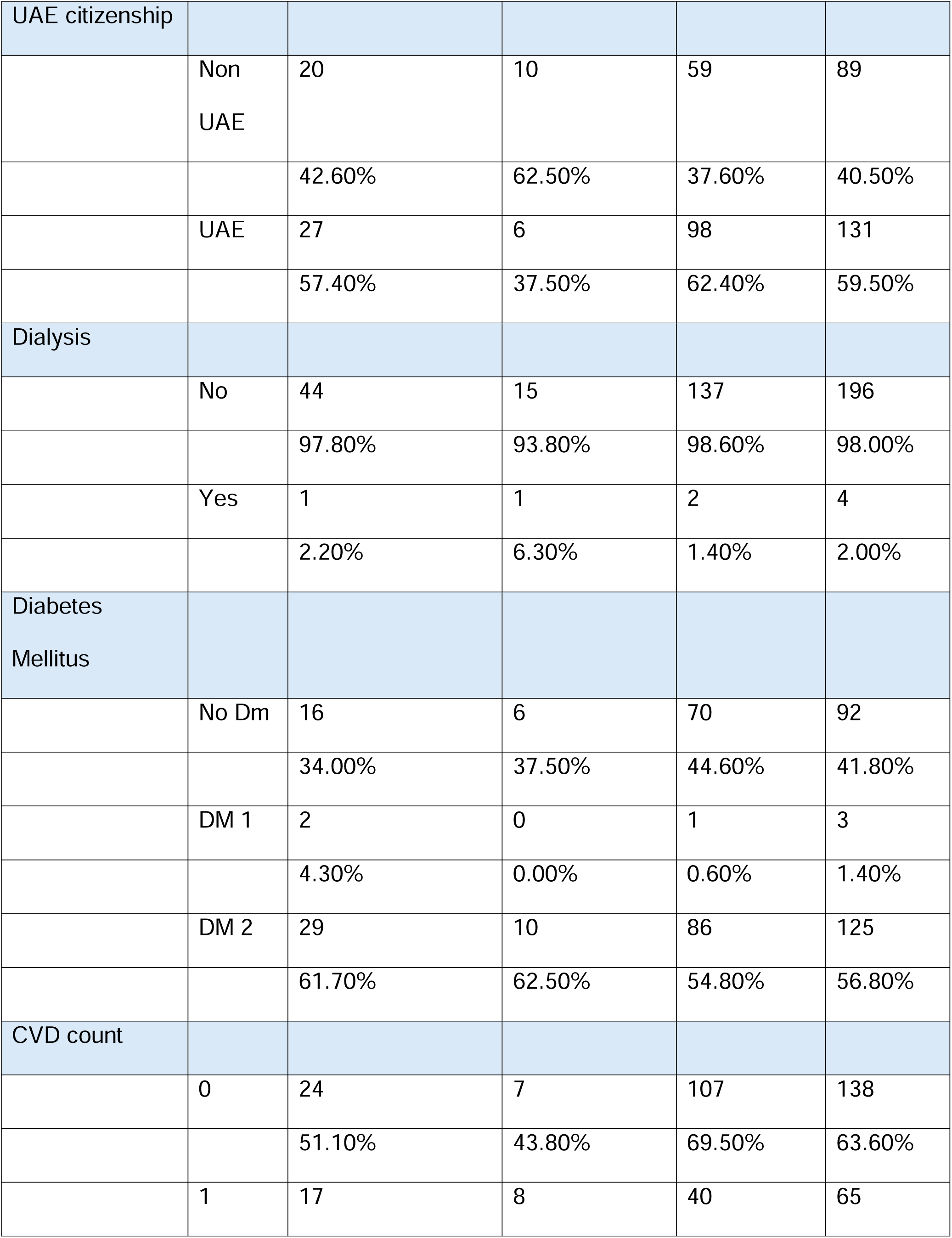
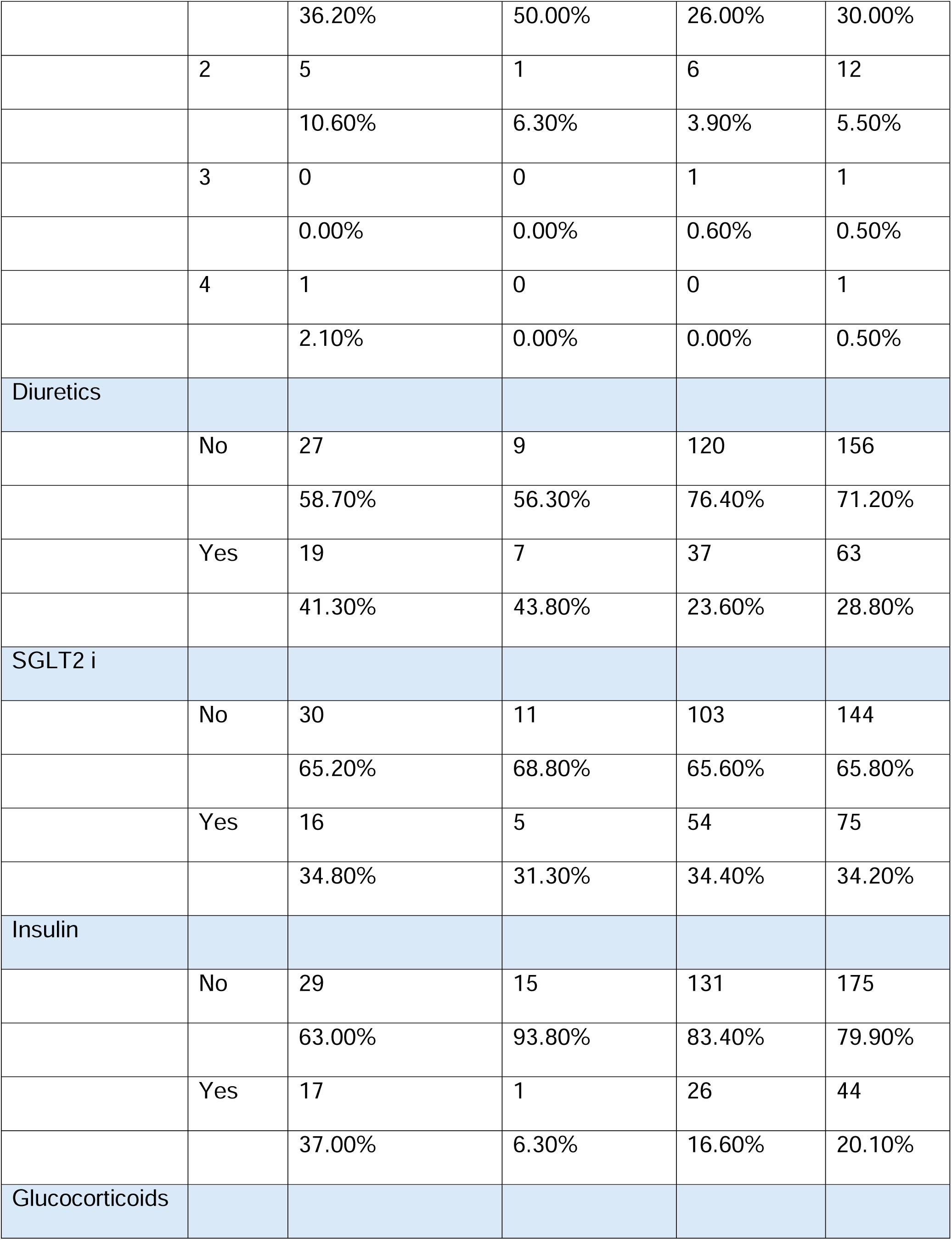
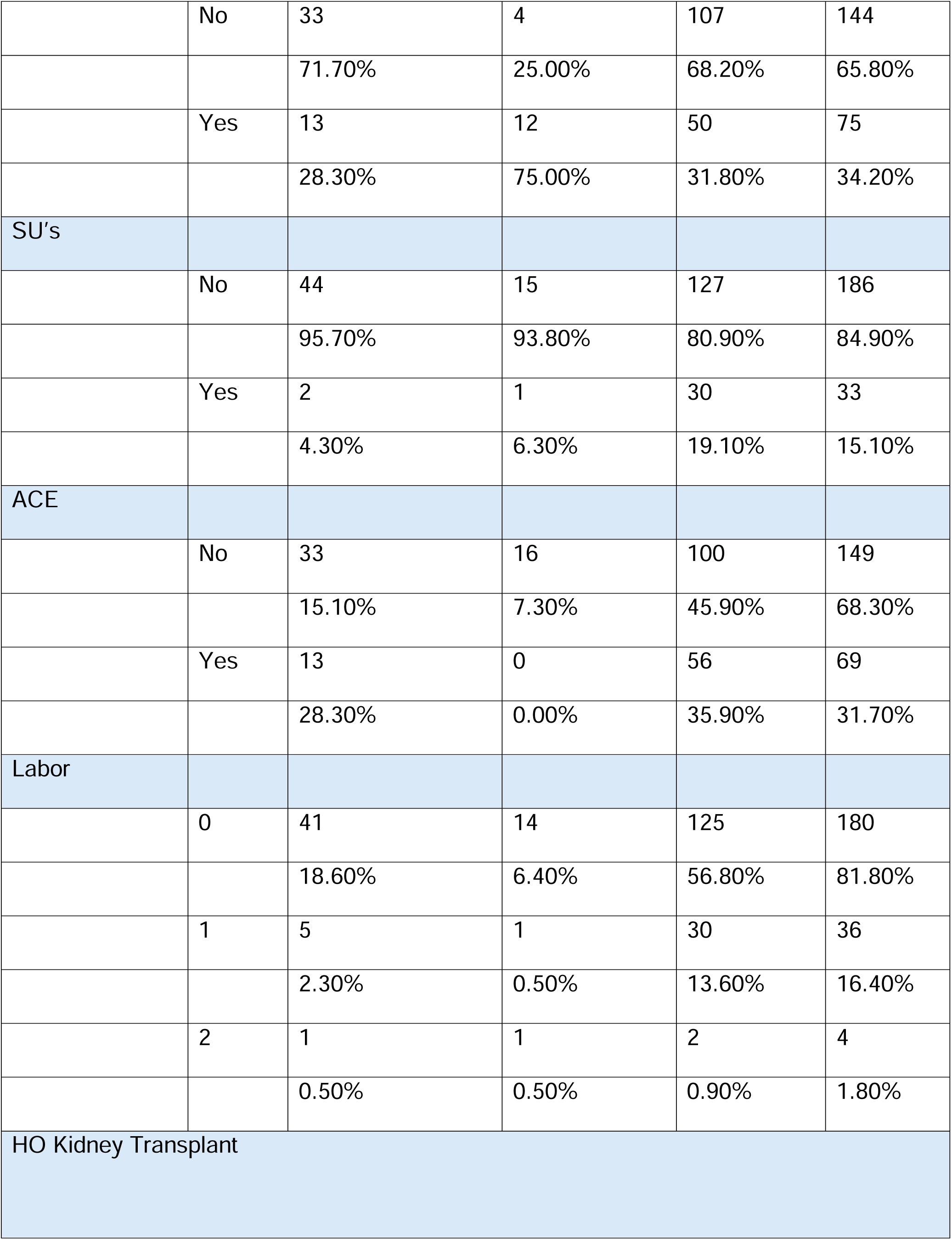
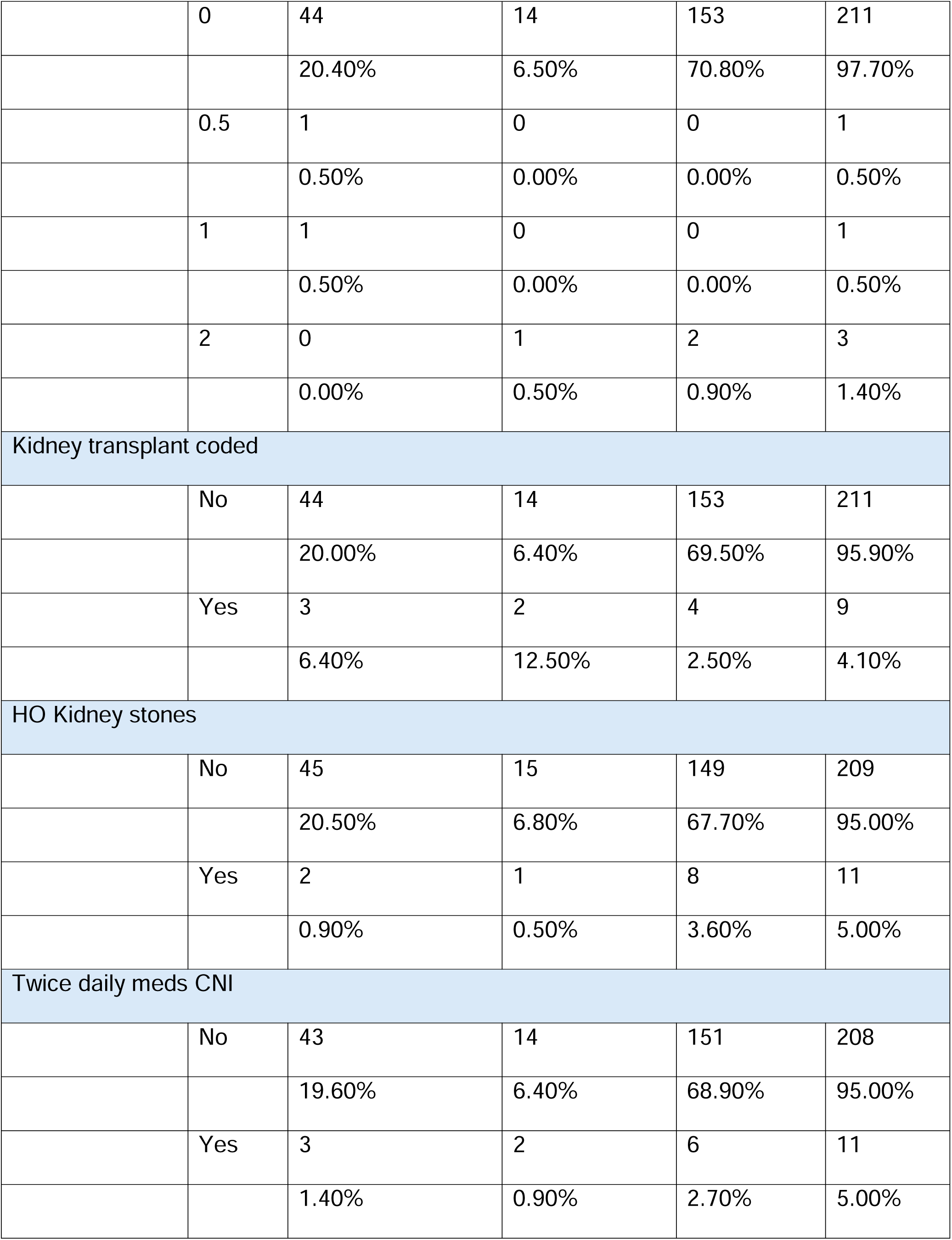
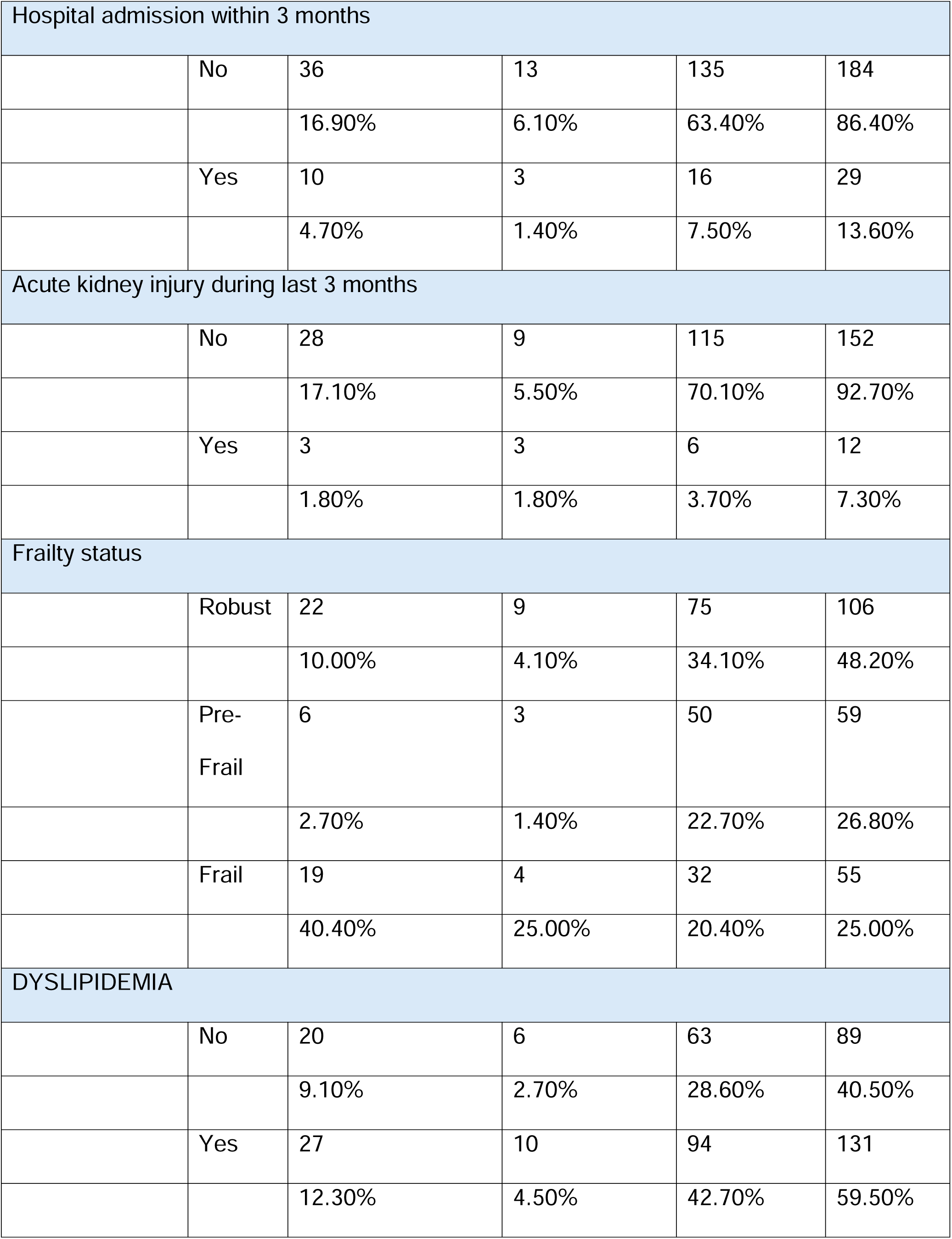
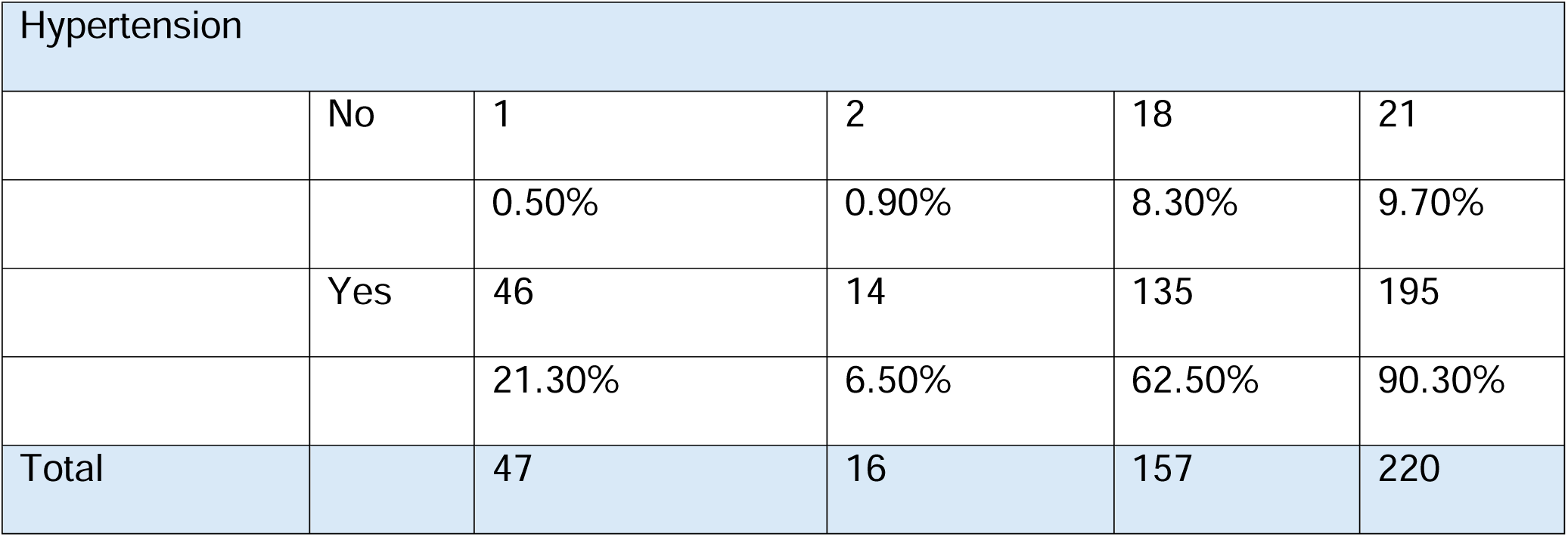
Subjects characteristics distributed by the fasting status.

This sample showed a mean RaK score of 9.78 and a median of 9; 55.5% had a RaK score of 9 or less, 24.5% 10-13, and 18.6% more than 13.

Figure 1 shows the RaK score distribution among the sample studied in relation to Ramadan fasting status and subjects experiencing ASE. These two outcomes, shown in Table 2, were studied using regression analysis in relation to the RaKtool and fasting status. Ordinal regression was used to explore predictors of fasting status, grouped as did not fast any day (y=0), fasted some days (y=1), or fasted all days (y=3) (Table 3). Independent variables considered were all variables included in the study.

**Figure 1:**
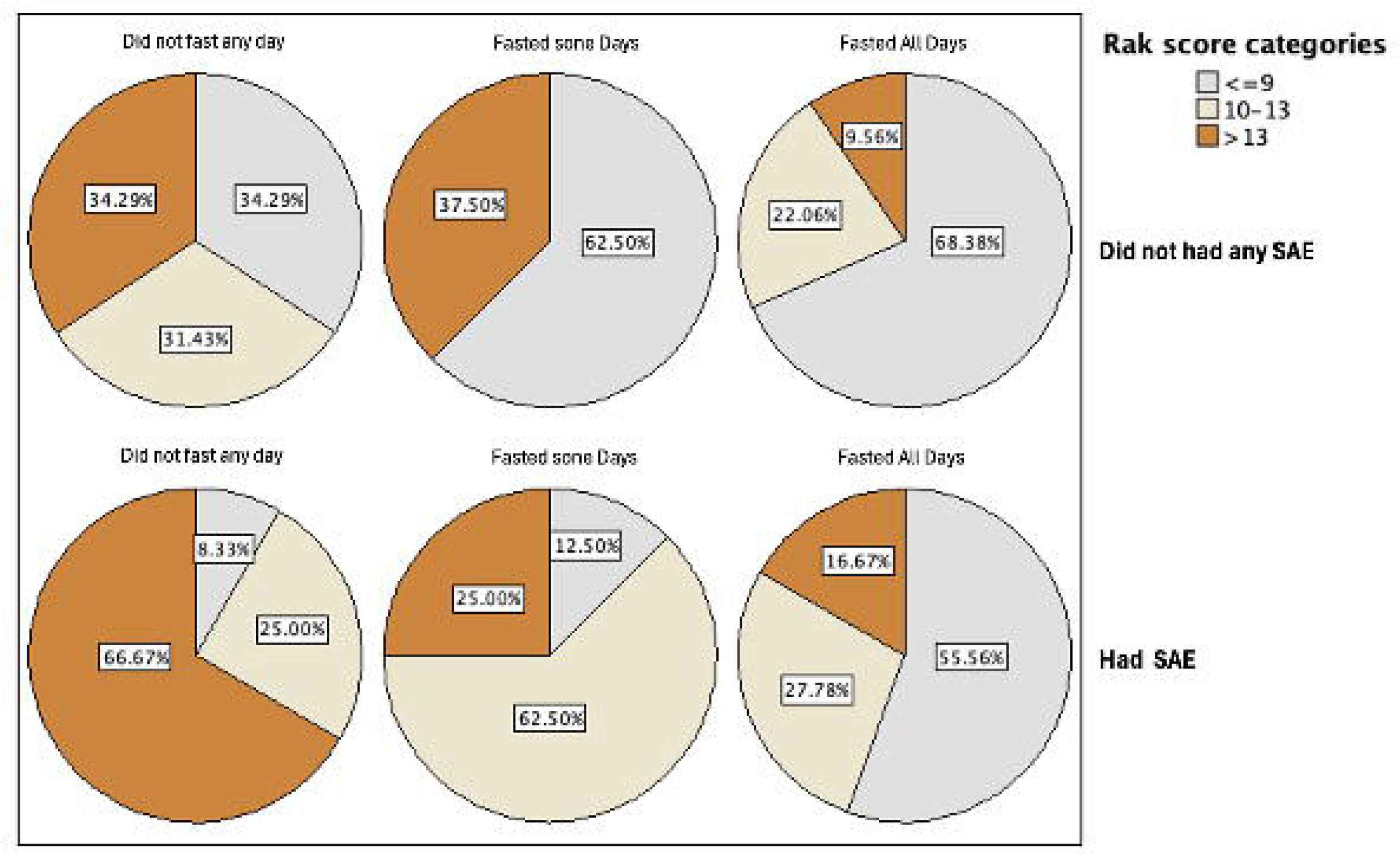
RaK score distribution by fasting status and occurrence of Significant Adverse Events.

**Table 2.**
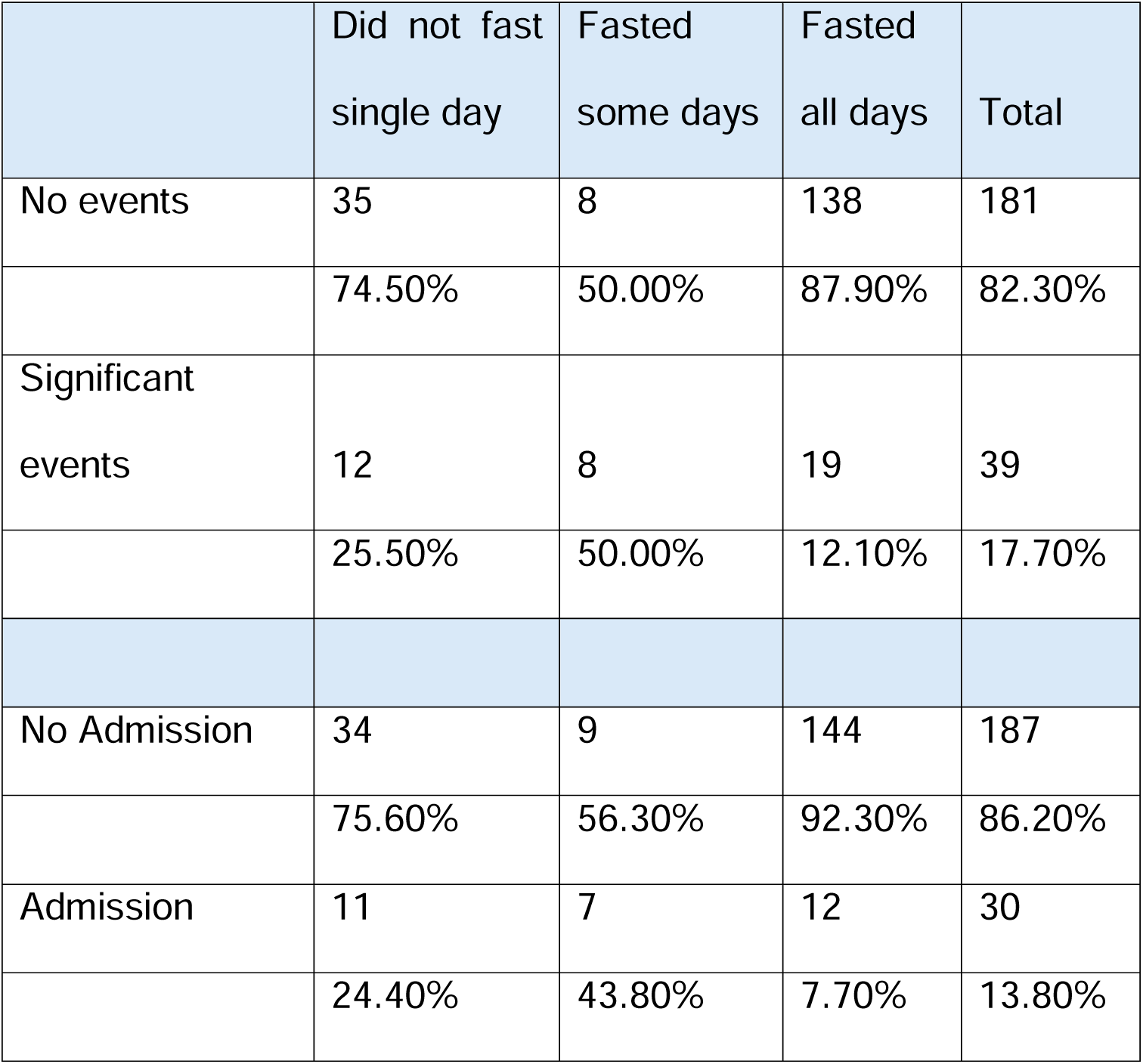
The relation between Fasting Ramadan status and the occurrence of Adverse Significant Event ASE.

**Table 3.**
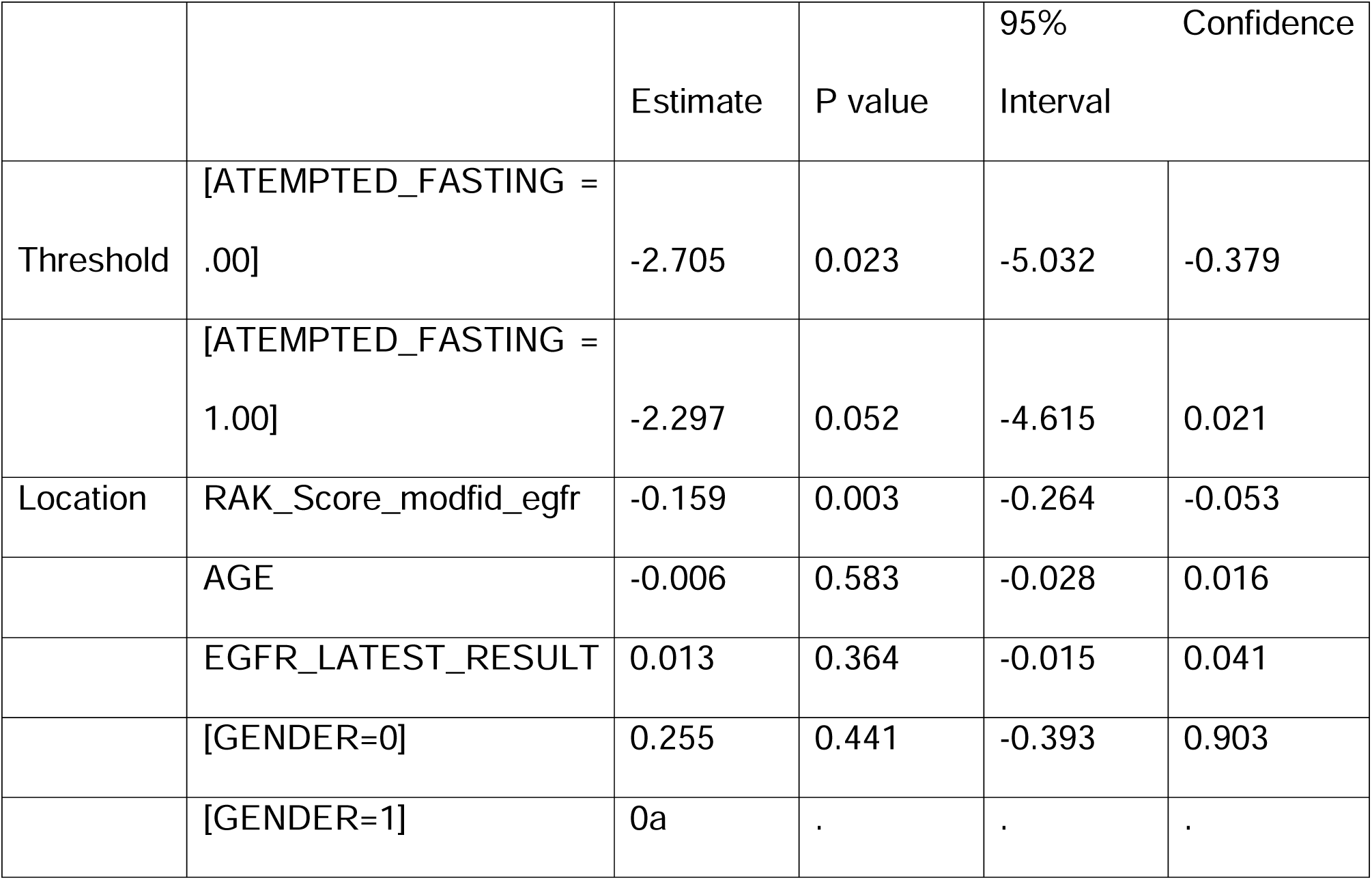
Prediction of the ability to fast using ordinal regression.

Only the RAK risk score was a significant predictor, with an estimate of −0.159 (−0.264 - −0.053), p-value=0.003. The AUC for predicting attempting fasting was 0.715 (0.635-0.794), with the best cutoff point of 9.5, a sensitivity of 69.8%, and a specificity of 66.9%. Figure 2.

**Figure 2:**
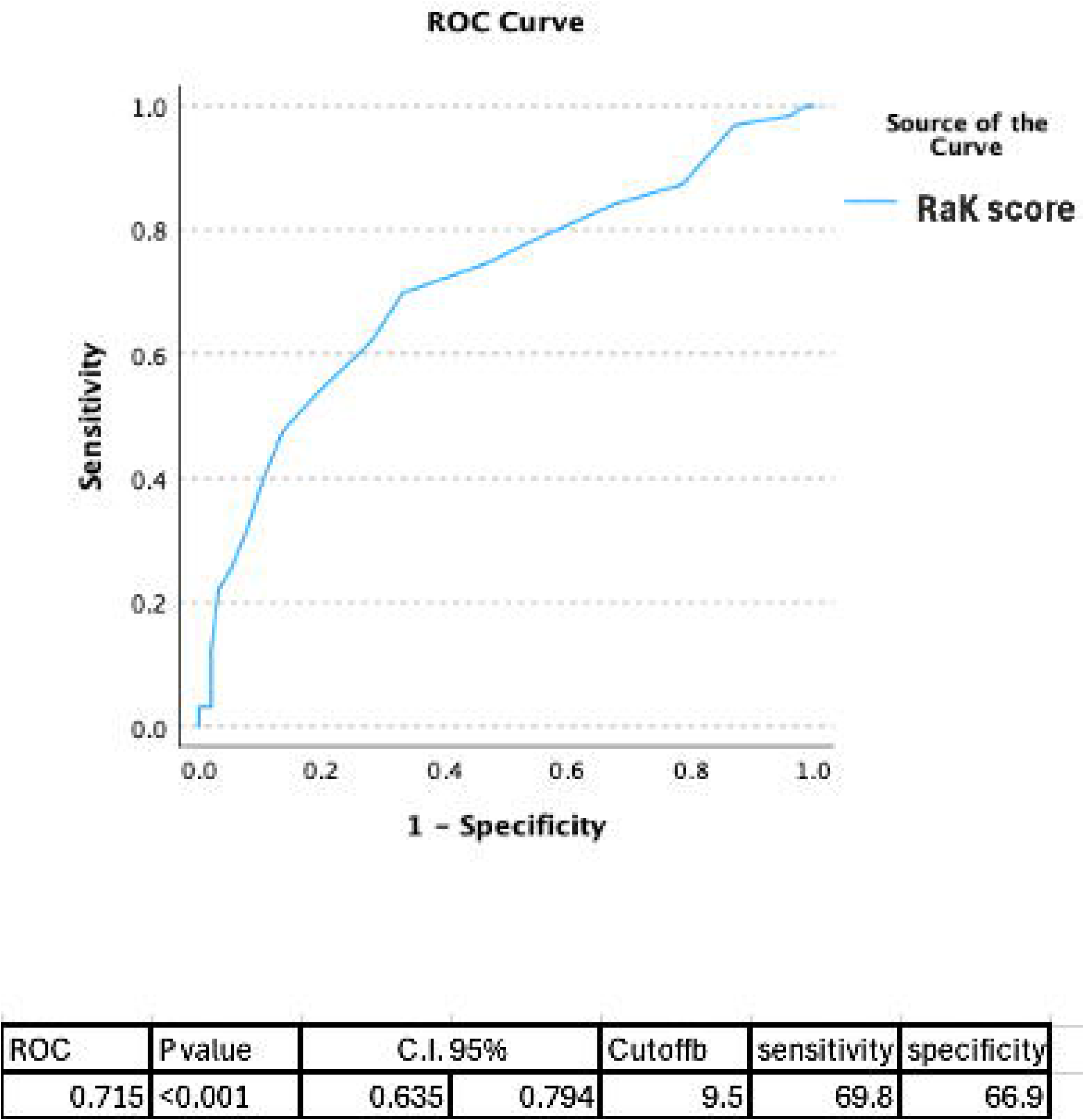
Area Under the Curve of the developed model.

When predicting SAE using logistic regression, a higher RAK score was significantly associated with the occurrence of events, with B= 0.253 (1.036-1.599) p-value= 0.023. Also, non-UAE nationality and frailty were associated with higher SAE (Table 3). The AUC to predict SAE was 0.689 (0.602-0.775), with a best cutoff point of 8.5, a sensitivity of 82.5%, and a specificity of 49.8%.

This ability of the RaK score to predict fasting status and SAEs is evident. All those with SAE who did not fast had a RaK score of 9 or above, and among those who had to break their fast (fasted at least a day), 7 out of the 8 (87.5%) had a score of 9 or more. Of those who had an SAE and completed all fasting days, 55.9 % had a RaK score of 9 or less, and only 16.7% had a score of 13 or more. Of patients who did not have any SAEs and completed fasting, 68.4% had a RaK score of 9 or less, and only 9.6% had a score of more than 13. Additionally, in the group of 8 patients who did not have any SAE but only fasted some days, 5 (62.5%) had a score of 9 or less, while those who did not fast, 34.3%, had a score of 9 or less. Figure 3

An important component of the RaK score is the eGFR. It performed relatively well in predicting fasting status, with an AUC of 0.677 (0.593 – 0.673), the eGFR however had a poor performance in predicting ASE, with an AUC of 0.590 (0.49-0.691).

**Figure 3:**
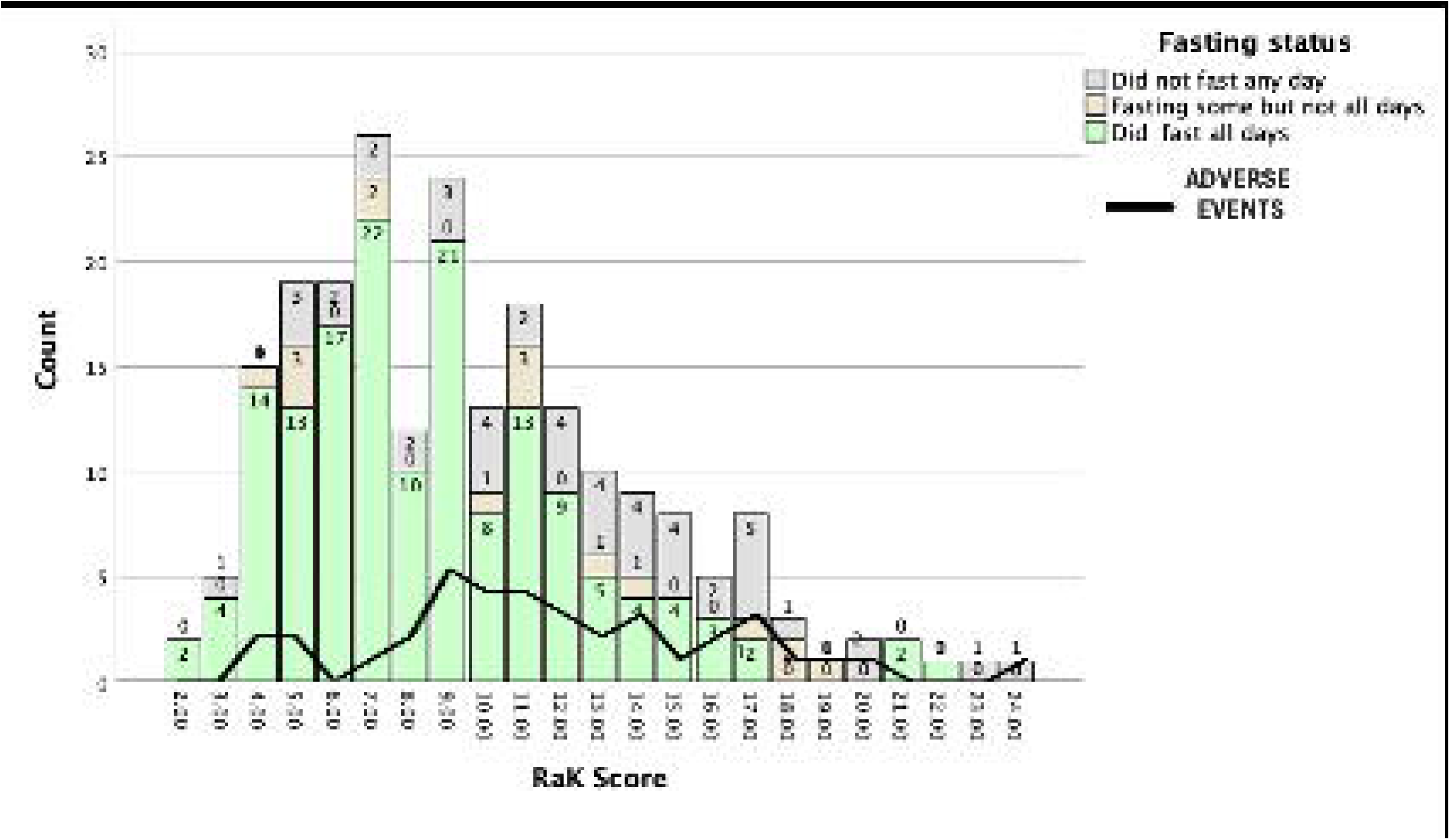
RaK score categories among patients who had Significant Adverse Events and those who did not distributed by fasting status.

**Table 4.**
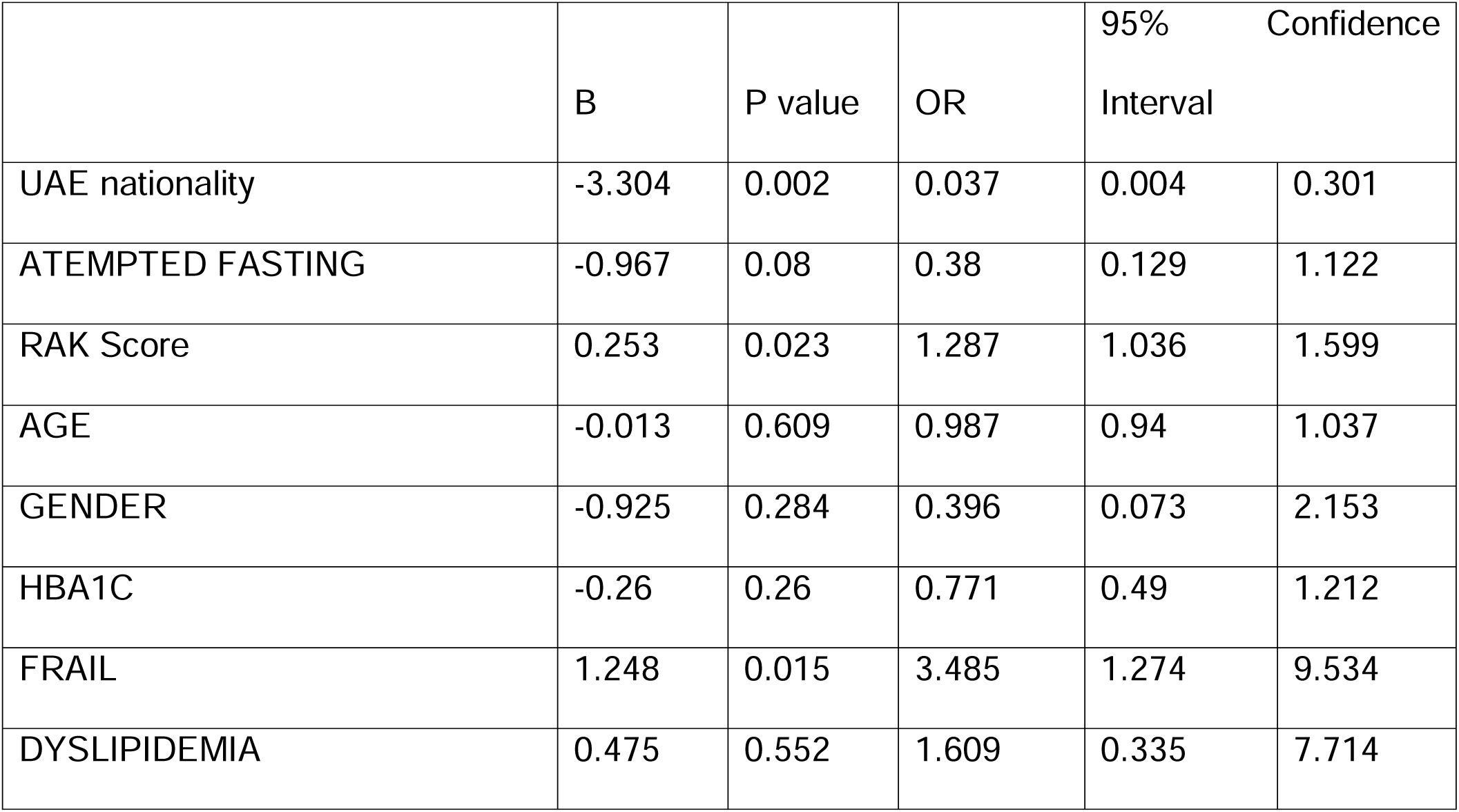
Risk factors for SAE among subjects who did fast at least one day of Ramadan.

## Discussion

This cohort had a strong representation of high-risk patients in an outpatient setting, including individuals with cardiac diseases, diabetes, hypertension, Proteinuria, and other comorbidities. This diversity enhances both the internal and external validity of our study. The RaK score effectively predicted both the ability to fast and the risk of significant adverse events (SAEs) among participants. This is the first study to validate this tool, but further research is needed to assess its applicability in special risk groups and larger, more heterogeneous samples. For instance, the growing number of dialysis patients and kidney transplant recipients who wish to fast represents a crucial area for further investigation.

Currently, the IDF-DAR tool is the only validated risk assessment tool for Ramadan fasting. Given that nearly half of the participants in our study had diabetes, the RaK tool could be used in conjunction with the IDF-DAR tool for CKD patients with diabetes who plan to fast.

The ability to fast is closely tied to a patient’s overall health and well-being, making it a valuable outcome measure for validation. Statistical evidence suggests that a RaK score of ≤9 indicates low risk, 10–13 indicates moderate risk, and>13 indicates high risk for SAEs. Specifically, categorization was based on the observation that only 9.6% of patients with a RaK score >13 fasted throughout Ramadan without experiencing any SAEs, while 68.4% of patients who completed fasting without SAEs had a RaK score of 9 or lower. Beyond quantifiable risk, an essential factor in decision-making is the acceptable level of harm as perceived by both healthcare providers and patients. Therefore, lower cutoff points for lower risk can be more considered after incorporating other factors related to patients’ social determinants of health and healthcare system resources. Such factors could explain UAE nationals experiencing fewer SAEs, which may reflect the influence of social determinants of health, such as work conditions, family support, health literacy, and financial status, an area that warrants further research.

The occurrence of SAEs was significantly associated with frailty status and non-UAE nationality. Similar studies (8) (9) (10)have also highlighted the role of frailty, emphasizing the need to strengthen the healthcare system with interventions and care guidelines focused on early detection and management of frailty. Other assessments of older patients can be assessed in future studies, such as Basic (BADL) and Instrumental (IADL) Activities of Daily Living scores. (11)

Although eGFR performed relatively well in predicting fasting status, it was less effective in predicting SAEs, which may be due to a bias in which physicians advise patients with lower eGFR not to fast. However, confirming this hypothesis is challenging, as randomization would be unethical, and patient preferences and adherence to medical advice are challenging to control.

Implementing this tool has implications that extend beyond improving care outcomes; it also supports physicians’ decision-making, enhances their knowledge, and promotes greater patient engagement in their care.

This study’s strengths include its prospective design and community-based sample. However, its limitations include insufficient power to assess the adequacy of the score for specific risk groups, such as transplant and dialysis patients. Additionally, the introduction of new medications may alter the clinical landscape, making future updates to the tool essential.

## Conclusion

The RaK risk assessment tool is a valid predictor of fasting status and significant adverse events (SAEs) in CKD patients. This validation is the first of a risk assessment tool designed for CKD patients intending to fast during Ramadan. Further studies are needed to refine its applicability and provide more tailored guidance for specific subgroups of kidney patients.

## Supporting information

RAK tool

## Data Availability

Data access is restricted as it is subject to the institution's data access policy.

## Declarations

### Ethical approval and consent to participate

The study was approved by the Ambulatory Healthcare Services IRB, approval number 2024-5. Consent statement in the Ethics approval and consent to participate: Informed consent was gathered verbally during the two occasions of calling patients before and after Ramadan.

## Acknowledgments

No funding was used in this study. Consent to publish: Not Applicable.

## Availability of data and materials

Data access is restricted as it is subject to the institution’s data access policy.

## Authors’ contributions

LBK and YB conceptualized the study. LBK and NN analyzed the data. LBK wrote the manuscript. EM, FM, MY, JS, AK, AA, AB, AS, SS, RK, MA, FK, AA, TF, WA, NY, HA collected data. All authors reviewed and approved the final manuscript.

