## Supplementary material for "Ramadan and Kidney disease (RaK) risk assessment tool. Potential Risk Calculator for Evaluating the Risk of Ramadan Fasting In Chronic Kidney Disease patients": Table 1

Table 1 Subjects characteristics distributed by the fasting status

|  |  | Did not fast single day | Fasted some days | Fasted all days | Total |
| --- | --- | --- | --- | --- | --- |
| Age groups | <=30 | 0 | 0 | 4 | 4 |
|  |  | 0.00% | 0.00% | 2.50% | 1.80% |
|  | 31-40 | 6 | 2 | 8 | 16 |
|  |  | 12.80% | 12.50% | 5.10% | 7.30% |
|  | 41-50 | 3 | 0 | 19 | 22 |
|  |  | 6.40% | 0.00% | 12.10% | 10.00% |
|  | 51-60 | 5 | 1 | 24 | 30 |
|  |  | 10.60% | 6.30% | 15.30% | 13.60% |
|  | 61-70 | 7 | 4 | 41 | 52 |
|  |  | 14.90% | 25.00% | 26.10% | 23.60% |
|  | 70-80 | 15 | 6 | 41 | 62 |
|  |  | 31.90% | 37.50% | 26.10% | 28.20% |
|  | >80 | 11 | 3 | 20 | 34 |
|  |  | 23.40% | 18.80% | 12.70% | 15.50% |
| Sex |  | 0 | 1 | 2 |  |
|  | Female | 25 | 6 | 83 | 114 |
|  |  | 53.20% | 37.50% | 52.90% | 51.80% |
|  | Male | 22 | 10 | 74 | 106 |
|  |  | 46.80% | 62.50% | 47.10% | 48.20% |
| UAE citizenship | 0 | 20 | 10 | 59 | 89 |
|  | Non UAE | 20 | 10 | 59 | 89 |
|  |  | 42.60% | 62.50% | 37.60% | 40.50% |
|  | UAE | 27 | 6 | 98 | 131 |
|  |  | 57.40% | 37.50% | 62.40% | 59.50% |
| Dialysis |  | 0 | 1 | 2 |  |
|  | No | 44 | 15 | 137 | 196 |
|  |  | 97.80% | 93.80% | 98.60% | 98.00% |
|  | Yes | 1 | 1 | 2 | 4 |
|  |  | 2.20% | 6.30% | 1.40% | 2.00% |
| dm012 |  | 0 | 1 | 2 |  |
|  | No Dm | 16 | 6 | 70 | 92 |
|  |  | 34.00% | 37.50% | 44.60% | 41.80% |
|  | DM 1 | 2 | 0 | 1 | 3 |
|  |  | 4.30% | 0.00% | 0.60% | 1.40% |
|  | DM 2 | 29 | 10 | 86 | 125 |
|  |  | 61.70% | 62.50% | 54.80% | 56.80% |
| CVD count |  | 0 | 1 | 2 |  |
|  | 0 | 24 | 7 | 107 | 138 |
|  |  | 51.10% | 43.80% | 69.50% | 63.60% |
|  | 1 | 17 | 8 | 40 | 65 |
|  |  | 36.20% | 50.00% | 26.00% | 30.00% |
|  | 2 | 5 | 1 | 6 | 12 |
|  |  | 10.60% | 6.30% | 3.90% | 5.50% |
|  | 3 | 0 | 0 | 1 | 1 |
|  |  | 0.00% | 0.00% | 0.60% | 0.50% |
|  | 4 | 1 | 0 | 0 | 1 |
|  |  | 2.10% | 0.00% | 0.00% | 0.50% |
| Diuretics |  | 0 | 1 | 2 |  |
|  | No | 27 | 9 | 120 | 156 |
|  |  | 58.70% | 56.30% | 76.40% | 71.20% |
|  | Yes | 19 | 7 | 37 | 63 |
|  |  | 41.30% | 43.80% | 23.60% | 28.80% |
| SGLT2 i |  | 0 | 1 | 2 |  |
|  | No | 30 | 11 | 103 | 144 |
|  |  | 65.20% | 68.80% | 65.60% | 65.80% |
|  | Yes | 16 | 5 | 54 | 75 |
|  |  | 34.80% | 31.30% | 34.40% | 34.20% |
| Insulin |  | 0 | 1 | 2 |  |
|  | No | 29 | 15 | 131 | 175 |
|  |  | 63.00% | 93.80% | 83.40% | 79.90% |
|  | Yes | 17 | 1 | 26 | 44 |
|  |  | 37.00% | 6.30% | 16.60% | 20.10% |
| Glucocorticoids |  | 0 | 1 | 2 |  |
|  | No | 33 | 4 | 107 | 144 |
|  |  | 71.70% | 25.00% | 68.20% | 65.80% |
|  | Yes | 13 | 12 | 50 | 75 |
|  |  | 28.30% | 75.00% | 31.80% | 34.20% |
| SU’s |  | 0 | 1 | 2 |  |
|  | No | 44 | 15 | 127 | 186 |
|  |  | 95.70% | 93.80% | 80.90% | 84.90% |
|  | Yes | 2 | 1 | 30 | 33 |
|  |  | 4.30% | 6.30% | 19.10% | 15.10% |
| ACE, |  | 0 | 1 | 2 |  |
|  | No | 33 | 16 | 100 | 149 |
|  |  | 15.10% | 7.30% | 45.90% | 68.30% |
|  | Yes | 13 | 0 | 56 | 69 |
|  |  | 28.30% | 0.00% | 35.90% | 31.70% |
| Labor |  | 0 | 1 | 2 |  |
|  | 0 | 41 | 14 | 125 | 180 |
|  |  | 18.60% | 6.40% | 56.80% | 81.80% |
|  | 1 | 5 | 1 | 30 | 36 |
|  |  | 2.30% | 0.50% | 13.60% | 16.40% |
|  | 2 | 1 | 1 | 2 | 4 |
|  |  | 0.50% | 0.50% | 0.90% | 1.80% |
| HO Kidney Transplant |  | 0 | 1 | 2 |  |
|  | 0 | 44 | 14 | 153 | 211 |
|  |  | 20.40% | 6.50% | 70.80% | 97.70% |
|  | 0.5 | 1 | 0 | 0 | 1 |
|  |  | 0.50% | 0.00% | 0.00% | 0.50% |
|  | 1 | 1 | 0 | 0 | 1 |
|  |  | 0.50% | 0.00% | 0.00% | 0.50% |
|  | 2 | 0 | 1 | 2 | 3 |
|  |  | 0.00% | 0.50% | 0.90% | 1.40% |
| Kidney transplant coded |  | 0 | 1 | 2 |  |
|  | No | 44 | 14 | 153 | 211 |
|  |  | 20.00% | 6.40% | 69.50% | 95.90% |
|  | Yes | 3 | 2 | 4 | 9 |
|  |  | 6.40% | 12.50% | 2.50% | 4.10% |
| HO Kidney stones |  | 0 | 1 | 2 |  |
|  | No | 45 | 15 | 149 | 209 |
|  |  | 20.50% | 6.80% | 67.70% | 95.00% |
|  | Yes | 2 | 1 | 8 | 11 |
|  |  | 0.90% | 0.50% | 3.60% | 5.00% |
| Twice daily meds CNI |  | 0 | 1 | 2 |  |
|  | No | 43 | 14 | 151 | 208 |
|  |  | 19.60% | 6.40% | 68.90% | 95.00% |
|  | Yes | 3 | 2 | 6 | 11 |
|  |  | 1.40% | 0.90% | 2.70% | 5.00% |
| Hospital admission within 3 months |  | 0 | 1 | 2 |  |
|  | No | 36 | 13 | 135 | 184 |
|  |  | 16.90% | 6.10% | 63.40% | 86.40% |
|  | Yes | 10 | 3 | 16 | 29 |
|  |  | 4.70% | 1.40% | 7.50% | 13.60% |
| Acute kidney injury during last 3 months |  | 0 | 1 | 2 |  |
|  | No | 28 | 9 | 115 | 152 |
|  |  | 17.10% | 5.50% | 70.10% | 92.70% |
|  | Yes | 3 | 3 | 6 | 12 |
|  |  | 1.80% | 1.80% | 3.70% | 7.30% |
| Frailty status |  | 0 | 1 | 2 |  |
|  | Robust | 22 | 9 | 75 | 106 |
|  |  | 10.00% | 4.10% | 34.10% | 48.20% |
|  | Pre-Frail | 6 | 3 | 50 | 59 |
|  |  | 2.70% | 1.40% | 22.70% | 26.80% |
|  | Frail | 19 | 4 | 32 | 55 |
|  |  | 40.40% | 25.00% | 20.40% | 25.00% |
| DYSLIPIDEMIA |  | 0 | 1 | 2 |  |
|  | No | 20 | 6 | 63 | 89 |
|  |  | 9.10% | 2.70% | 28.60% | 40.50% |
|  | Yes | 27 | 10 | 94 | 131 |
|  |  | 12.30% | 4.50% | 42.70% | 59.50% |
| Hypertension |  | 0 | 1 | 2 |  |
|  | No | 1 | 2 | 18 | 21 |
|  |  | 0.50% | 0.90% | 8.30% | 9.70% |
|  | Yes | 46 | 14 | 135 | 195 |
|  |  | 21.30% | 6.50% | 62.50% | 90.30% |
| Total |  | 47 | 16 | 157 | 220 |
