## Supplementary material for "Ramadan and Kidney disease (RaK) risk assessment tool. Potential Risk Calculator for Evaluating the Risk of Ramadan Fasting In Chronic Kidney Disease patients": Table 2

Table 2 The relation between Fasting Ramadan status and the occurrence of Adverse Significant Event ASE.

|  | Did not fast single day | Fasted some days | Fasted all days | Total |
| --- | --- | --- | --- | --- |
| No events | 35 | 8 | 138 | 181 |
|  | 74.50% | 50.00% | 87.90% | 82.30% |
| Significant events | 12 | 8 | 19 | 39 |
|  | 25.50% | 50.00% | 12.10% | 17.70% |
|  | 0 | 1 | 2 |  |
| No Admission | 34 | 9 | 144 | 187 |
|  | 75.60% | 56.30% | 92.30% | 86.20% |
| Admission | 11 | 7 | 12 | 30 |
|  | 24.40% | 43.80% | 7.70% | 13.80% |
