## Supplementary material for "Ramadan and Kidney disease (RaK) risk assessment tool. Potential Risk Calculator for Evaluating the Risk of Ramadan Fasting In Chronic Kidney Disease patients": Table 3

Table 3 Prediction of the ability to fast using ordinal regression

|  |  | Estimate | P value | 95% Confidence Interval | |
| --- | --- | --- | --- | --- | --- |
| Threshold | [ATEMPTED_FASTING = .00] | -2.705 | 0.023 | -5.032 | -0.379 |
|  | [ATEMPTED_FASTING = 1.00] | -2.297 | 0.052 | -4.615 | 0.021 |
| Location | RAK_Score_modfid_egfr | -0.159 | 0.003 | -0.264 | -0.053 |
|  | AGE | -0.006 | 0.583 | -0.028 | 0.016 |
|  | EGFR_LATEST_RESULT | 0.013 | 0.364 | -0.015 | 0.041 |
|  | [GENDER=0] | 0.255 | 0.441 | -0.393 | 0.903 |
|  | [GENDER=1] | 0a | . | . | . |
