## Supplementary material for "Ramadan and Kidney disease (RaK) risk assessment tool. Potential Risk Calculator for Evaluating the Risk of Ramadan Fasting In Chronic Kidney Disease patients": Table 4

Table 4 Risk factors for SAE among subjects who did fast at least one day of Ramadan

|  | B | P value | OR | 95% C.I. | |
| --- | --- | --- | --- | --- | --- |
| UAE nationality | -3.304 | 0.002 | 0.037 | 0.004 | 0.301 |
| ATEMPTED FASTING | -0.967 | 0.08 | 0.38 | 0.129 | 1.122 |
| RAK Score | 0.253 | 0.023 | 1.287 | 1.036 | 1.599 |
| AGE | -0.013 | 0.609 | 0.987 | 0.94 | 1.037 |
| GENDER | -0.925 | 0.284 | 0.396 | 0.073 | 2.153 |
| HBA1C | -0.26 | 0.26 | 0.771 | 0.49 | 1.212 |
| FRAIL | 1.248 | 0.015 | 3.485 | 1.274 | 9.534 |
| DYSLIPIDEMIA | 0.475 | 0.552 | 1.609 | 0.335 | 7.714 |
