## Supplementary material for "Ramadan and Kidney disease (RaK) risk assessment tool. Potential Risk Calculator for Evaluating the Risk of Ramadan Fasting In Chronic Kidney Disease patients": RAK tool

### Risk Calculation for Ramadan Fasting and Kidney Diseases (RaK) 2022

| Potential Risk Factors | Projected Score | Total Score |
|---|---|---|
| 1. Age<br>1. <70<br>2. >70 | <br>0<br>1 | |
| 2. GFR<br>1. < 60 mL/min/m2<br>2. < 45 mL/min/m2<br>3. < 30 mL/min/m2<br>4. < 15 mL/min/1.73 m2<br>5. On dialysis | <br>1<br>2<br>3<br>4<br>5 | |
| 3. Dialysis duration < 5 years | 1 | |
| 4. level of Proteinuria<br>1. No proteinuria<br>2. 0- <1 g/D<br>3. 1-3.5 g/D<br>4. >3.5 g/D | <br>0<br>0<br>1<br>2 | |
| 5. Presence of DM<br>1. No DM<br>2. T2DM<br>3. T1 DM | <br>0<br>1<br>2 | |
| 6. Glycemic Control<br>1. A1c <7.0<br>2. A1c 7-9%<br>3. A1c>9 | <br>0<br>1<br>2 | |
| 4. HTN<br>1. No<br>2. Well controlled<br>3. Poorly controlled | <br>0<br>0<br>2 | |
| 5. Presence of CVD<br>1. No<br>2. Stable MVD<br>3. Unstable MVD<br>4. Compensated Congestive heart failure<br>5. Cardiac arrhythmia | <br>0<br>1<br>2<br>1<br><br>1 | Adding 1 for any heart diseases if stable. If not stable add another 2 points |
| 6. HO Kidney Transplant<br>1. No<br>2. More than 12 months<br>3. Within 6-12 months<br>4. Within 1-6 months | <br>0<br>1<br>2<br>3 | |
| 7. HO Kidney stones<br>1. No<br>2. Yes | <br>0<br>1 | |
| 8. Medications<br>1. Twice daily meds CNI<br>2. Diuretics | <br>0<br>0 | Diuretics or SGLT2i or ACEI one point is added |

| | | |
|---|---|---|
| 3. SGLT2 i<br>4. ARB/ACE, I | 0<br>0 | |
| 9. Duration of fasting<br>1. < 16 hours<br>2. > 16 hours | <br>0<br>1 | |
| 10. patient expose to Temperature > 30<br>1. No<br>2. Yes | <br>0<br>1 | No air conditioning |
| 11. Labor<br>1. No physical labor<br>2. Moderate labor work<br>3. Heavy Physical labor | <br>0<br>1<br>2 | Labor during fasting |
| 12. Frailty and Cognitive Function<br>1. No frailty or loss in cognitive function | <br>0 1 2 | FRAIL score added |
| 13. Past fasting experience<br>1. No adverse events<br>2. Adverse events | <br>0<br>1 | |
| 14. Hospital admission within the last 3 months<br>1. Yes<br>2. No | <br>0<br>1 | |
| 15.Hx Acute kidney injury during last 3 months<br>1. Yes<br>2. No | <br><br><br>1<br>0 | |

**Risk Score and Risk Stratification**

| Risk Score | Risk level | Medical Recommendations | Religious Recommendations |
|---|---|---|---|
| <5 | Very Low Risk | **Fasting is probably safe**<br>1. Medical Evaluation<br>2. Medications adjustment<br>3. Strict monitoring<br>4. Check weight and GFR in 1 week | Fasting is obligatory |
| <9 | Low risk | | |
| **9-13** | **Moderate Risk** | **Fasting Safety is uncertain**<br>1. Medical Evaluation<br>2. Medications adjustment<br>3. Strict monitoring<br>4. Check weight and GFR in 1 week | 1. Fasting is preferred<br>2. patients may choose not to fast if concerned about their health after consulting the doctor and taking into account the full medical circumstances and patient own previous experience.<br>3. If patients fast, they must follow medical recommendations |
| **>13** | **High Risk** | **Fasting is probably unsafe**<br>1, Advise against fasting | Advise against fasting |
